# Factors predicting serum clozapine levels in Middle Eastern patients: an observational study

**DOI:** 10.1101/2022.01.02.22268620

**Authors:** Ahmed Hassab Errasoul, Mohammed A. Alarabi

**Author notes:** Corresponding author: Mohammed A. Alarabi, MBBS, Senior Registrar Psychiatrist, Department of Psychiatry, College of Medicine, King Saud University, PO Box 7805, Riyadh 11472, Kingdom of Saudi Arabia.

## Abstract

**Background:** Despite its superiority over other drugs for psychosis, clozapine remains underused and is associated with many clinical challenges, including difficulties in predicting serum levels. We found no large or recent study that investigated the determinants of serum clozapine levels in Middle Eastern patients. We thus investigated the association between clozapine dose and serum levels and the clinical predictors of clozapine serum levels in Middle Eastern patients.

**Methods:** This was a cross-sectional study including 94 patients of Middle Eastern ethnicity attending the Clozapine Clinic in King Saud University Medical City in Riyadh, Saudi Arabia.

**Results:** The average clozapine dose and serum level were 400 mg/daily and 705 ng/mL, respectively. Clozapine dose and serum levels were positively correlated (*r*_*s*_ [94]=0.32, *p*=0.002). We generated a predictive model of serum clozapine levels which revealed that the daily dose, smoking status, use of fluvoxamine or lamotrigine, and body mass index (BMI) predicted 43.6% of the variance in serum levels (*p*<0.001). Using this model, we calculated that the maximum doses of clozapine to avoid levels above the optimal range (>650 ng/mL) were 300, 250, or 225 mg/day for non-smokers with a body mass index of 25, 30, or 35 kg/m^2^, respectively, and 475, 425, or 400 mg/day for smokers with a body mass index of 25, 30, or 35 kg/m^2^, respectively.

**Conclusions:** This is a naturalistic study of the clozapine dose-level relationship and the clinical predictors of serum clozapine levels in Middle Eastern patients. These findings do not reduce the value of individualised therapeutic drug monitoring but may assist clinicians when prescribing clozapine to Middle Eastern patients. Further psychopharmacological studies are needed on this demographic population.

## Background

Clozapine has been demonstrated to be superior to other drugs for the treatment of schizophrenia symptoms [1-3]. Furthermore, it is the only approved medication for treatment-resistant schizophrenia [4]. However, clozapine remains underused in clinical practice [5], including in Saudi Arabia [6]. Furthermore, discontinuation of clozapine is common owing to nonadherence to monitoring protocols or drug-related side effects [7,8]. Such clinical challenges may be the reason behind the underutilisation of clozapine in Saudi Arabia and other Gulf countries [9]. One method of addressing these clinical concerns is therapeutic drug monitoring (TDM), which is the practice of measuring drug levels in the patient’s blood to adjust prescribed doses [10]. Studies have traditionally concluded that a serum clozapine level of 350 ng/mL, or a level that falls between 250 and 550 ng/mL, is required for clinical response [11-16]. Evidence suggests that serum levels exceeding 600–700 ng/mL are associated with lower rates of clinical responses and higher rates of dose-dependent side effects [17]. Remarkably, serum clozapine levels exhibit significant inter-individual variability [18], which may be related to ethnic differences in clozapine metabolism, and a resultant distinct relationship between clozapine dose and serum level in different ethnic groups. For instance, East Asians have been observed to require almost half the dose of clozapine to achieve the same serum levels as those in Caucasians of Anglo-Saxon lineage [19]. Moreover, findings from a study on 26 Middle Eastern patients from Saudi Arabia have suggested that Middle Eastern patients require doses lower than those required by European Caucasians to achieve the same therapeutic serum levels [13,19-22]. Our literature review found no large or recent study that investigated the determinants of serum clozapine levels in Middle Eastern patients. Therefore, the main aim of this study was to explore the association between clozapine dose and serum levels in an ethnically homogeneous group of Middle Eastern patients and to identify which clinical factors, besides daily clozapine dose, are associated with serum clozapine levels. We hypothesised that the association between clozapine dose and serum level in our patients would be linear as previously reported in other samples [13,20,23], and the most important predictors of variation in serum levels would be the daily dose of clozapine and the concomitant use of other psychotropic medications.

## Methods

This study was conducted at the Clozapine Clinic in King Khalid University Hospital (KKUH), part of King Saud University Medical City in Riyadh, Saudi Arabia. We included data of all patients attending the Clozapine Clinic in KKUH. The data encompassed the period between December 2015 (the inception of the Clozapine Clinic in KKUH) and April 2020. We collected data from patients’ electronic medical records and laboratory records. The inclusion criteria were clozapine treatment and at least one measurement of serum clozapine level. The exclusion criteria were undetectable serum clozapine level and missing data from electronic medical records regarding clozapine dose or concomitant psychotropic use. Collected data included demographics, psychiatric diagnosis, body mass index (BMI), and concomitant use of psychotropic medications other than clozapine. All included patients were Saudi nationals of Middle Eastern ethnicity. Blood samples were collected at KKUH approximately 12 h after the last oral dose of clozapine [24]. Samples were sent to the National Reference Laboratory in Abu Dhabi, United Arab Emirates, an outsourced laboratory service provider managed by LabCorp. The unit of serum level measurements was nanogram/millilitre, and levels above 650 ng/mL were considered above the optimal range [25]. For patients who had more than one measurement of serum clozapine levels, the first measurement was used in the analysis.

### Statistical Analysis

The Statistical Package for the Social Sciences (SPSS) version 21.0 [26] was used for data analysis. Descriptive data are presented as means (M) and standard deviations (SD) or medians (Mdn) and interquartile ranges (IQR). Correlations were estimated using bivariate correlation coefficients, Pearson’s coefficient (*r*) for normally distributed data, and non-parametric Spearman’s coefficient (*r*_*s*_) for non-normally distributed data. For normally distributed data, differences between groups were analysed using the Student’s t-test (*t*) or one-way analysis of variance (ANOVA) test (*F*) when appropriate. The Mann–Whitney U test (*U*) or Kruskal–Wallis one-way analysis of variance test (*H*) were used for non-normally distributed data. A hierarchical linear regression model was used to examine the predictors of serum clozapine levels in our sample. The level of significance (*p*) was fixed at <0.05 for analysis interpretation. The analysed dataset is available from the corresponding author on reasonable request.

### Ethical Standards

The study was approved by the Institutional Research Board Committee at the College of Medicine at King Saud University (approval reference no. 19/0054/IRB for Research Project No. E-19-4246). The requirement for informed consent was waived by the institutional review board because the study employed a retrospective observational design, and patients’ data remained confidential. The authors assert that all procedures contributing to this work comply with the ethical standards of the relevant national and institutional committee on human experimentation with the Declaration of Helsinki of 1975, as revised in 2008. The authors also followed the ICMJE guidelines in the composition of this manuscript.

## Results

Data of 94 patients attending the Clozapine Clinic were included in the analysis. Patients’ characteristics and primary psychiatric diagnoses are presented in Table 1. The mean BMI was 29.91 kg/m^2^ (standard deviation [*SD*] = 6.73, 95% confidence interval [CI]: 28.52– 31.29), ranging from 16.20 to 49.88 kg/m^2^, with half the sample (48.9%) having a BMI of 30 kg/m^2^ or above. Smoking was documented in 20 (21.3%) patients, 19 of whom were men.

**Table 1.** Patients’ characteristics.

|  | Men | Women | All patients |
| --- | --- | --- | --- |
| Median age (*IQR*) | 38 (30–47) | 34 (27–46) | 37 (29–46) |
| Schizophrenia (%) | 44 (46.8) | 24 (25.5) | 68 (72.3) |
| Schizoaffective, bipolar type (%) | 5 (5.3) | 3 (3.2) | 8 (8.5) |
| Schizoaffective, depressive type (%) | 3 (3.2) | 1 (1.1) | 4 (4.3) |
| Bipolar 1 disorder (%) | 3 (3.2) | 8 (8.5) | 11 (11.7) |
| Other diagnoses (%) | 2 (2.1) | 1 (1.1) | 3 (3.2) |
| Total (%): | 57 (60.6) | 37 (39.4) | 94 (100) |

IQR: interquartile range

The prescribed dose of clozapine ranged from 75 mg to 800 mg daily, with a mean daily dose of 400 mg (*SD* = 149.69, 95% CI: 369.34–430.66). No significant difference was found in the average daily dose of clozapine between male (*M* = 408.33 mg, *SD* = 149.28) and female patients (*M* = 387.16 mg, *SD* = 151.45) in our sample (*t* (92) = 0.67, *p* = 0.506, 95% CI: −41.78–84.12). Furthermore, no significant difference was observed in the average daily dose between patients who received additional psychotropic medications and those who received clozapine monotherapy (*p* > 0.150). The other psychotropic medications concomitantly used by patients during measurements of serum clozapine levels are presented in Table 2. In total, 64.9% of the patients in our sample were taking other psychotropics in addition to clozapine.

**Table 2.** Frequency of polytherapy during clozapine treatment.

| Drug class | Frequency (% of total sample) |
| --- | --- |
| Concomitant use of drugs for depression and anxiety | 38 (40.4) |
| Fluoxetine | 9 (9.6) |
| Escitalopram | 9 (9.6) |
| Fluvoxamine | 4 (4.3) |
| Sertraline | 2 (2.1) |
| Paroxetine | 2 (2.1) |
| Venlafaxine | 3 (3.2) |
| Imipramine | 1 (1.1) |
| Clomipramine | 1 (1.1) |
| Amitriptyline | 2 (2.1) |
| Mirtazapine | 5 (5.3) |
| Concomitant use of drugs for relapse prevention | 25 (25.6) |
| Valproate | 11 (11.7) |
| Lamotrigine | 3 (3.2) |
| Lithium | 9 (9.6) |
| Lithium and valproate | 1 (1.1) |
| Concomitant use of other drugs for psychosis | 14 (14.9) |
| Amisulpride | 3 (3.2) |
| Aripiprazole | 9 (9.6) |
| Quetiapine | 1 (1.1) |
| Risperidone | 1 (1.1) |

Our analysis showed that the median serum clozapine level was 705 ng/mL (interquartile range [*IQR*] = 442.25–979.50, 95% CI: 598.51–816). The majority of patients (54.3%) had serum levels higher than 650 ng/mL, which is considered above the optimal range as discussed previously. We found that serum clozapine levels and daily doses were positively correlated (*r*_*s*_ [94] = 0.32, *p* = 0.002, 95% CI: 0.13–0.49). The details of correlation analysis for patients on monotherapy and patients receiving other psychotropics are presented in Table 3. We also found that serum clozapine levels were higher in women (median [*Mdn*] = 843 ng/mL, *IQR* = 552–1048) than in men (*Mdn* = 607 ng/mL, *IQR* = 379–862), and this difference was significant (*U* = 768, *p* = 0.027). Non-smokers (*Mdn* = 766 ng/mL, *IQR* = 494–985.5) had significantly higher serum clozapine levels than smokers did (*Mdn* = 545 ng/mL, *IQR* = 209–843.25) (*U* = 520.5, *p* = 0.043). A trend for a positive correlation between measured clozapine levels and BMI was noted (*r*_*s*_ [93] = 0.20, *p* = 0.051, 95% CI: −0.01–0.39).

**Table 3.**
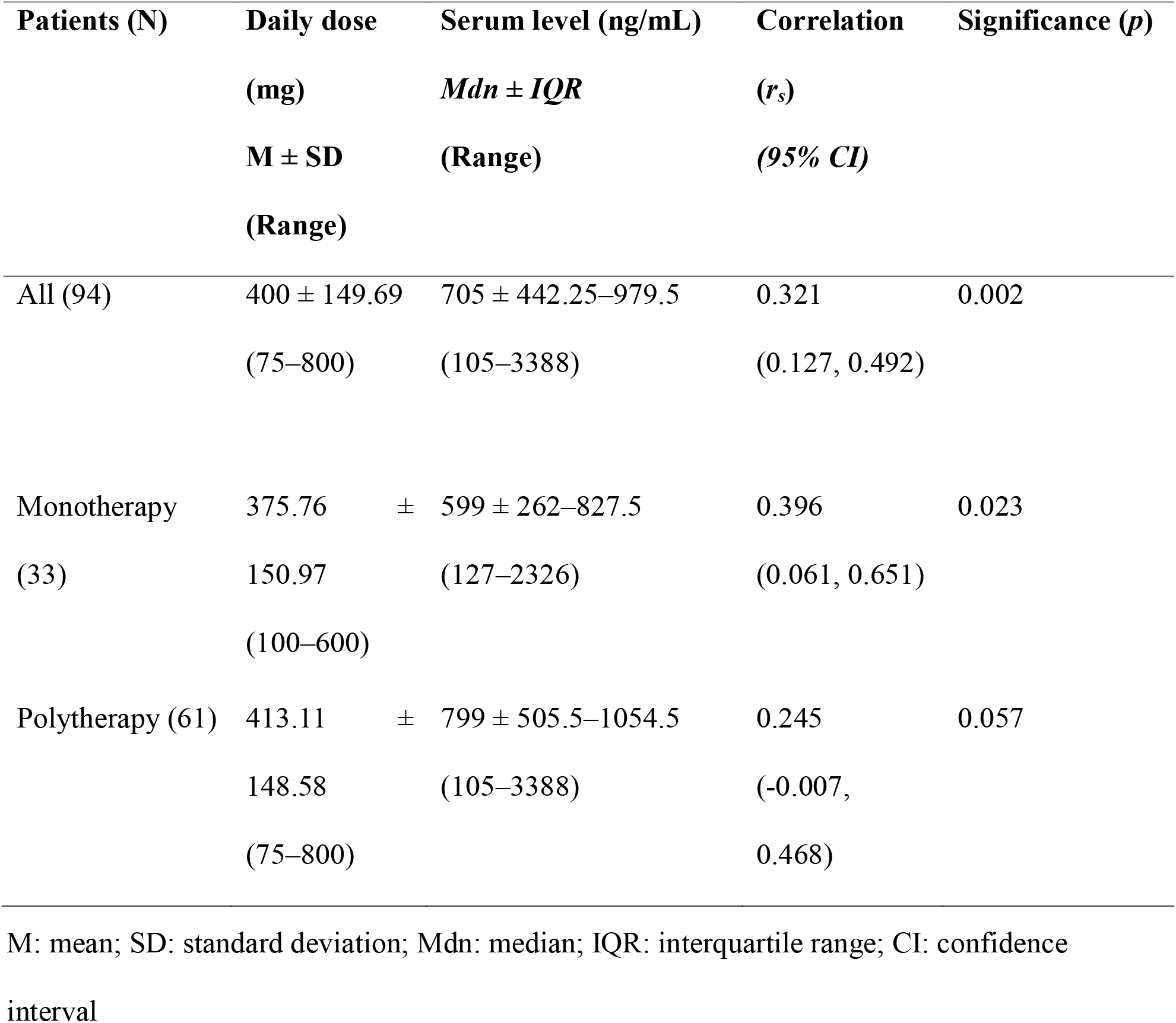
Correlation between daily clozapine dose and serum clozapine levels.

Patients receiving psychotropic medications other than clozapine had higher serum clozapine levels (*Mdn* = 799 ng/mL, *IQR* = 505.5–1054.5, 95% CI: 634–904) than patients receiving clozapine monotherapy (*Mdn* = 599, *IQR* = 262–827.5, 95% CI: 357.64–779), and this difference was significant (*U* = 714, *p* = 0.021). Among patients concomitantly receiving drugs for depression and anxiety (n = 38), patients receiving fluvoxamine (n = 4) had the highest average serum clozapine level (*Mdn* = 2733 ng/mL, *IQR* = 2373.25– 3248.75, 95% CI: 2286, 3388), which was significantly higher than the serum level of all other patients in our sample (*U* = 1.00, *p* < 0.001). In addition, those receiving lamotrigine (n = 3) had higher average serum clozapine levels (*Mdn* = 1457 ng/mL, *IQR* = 1344.5– 1810.5, 95% CI: 1232, 2164) than the rest of the sample (*U* = 19.00, *p* = 0.011). No significant differences were observed in serum clozapine levels between patients receiving other psychotropic medications and other patients not receiving those medications (*p* > 0.05).

Hierarchical multiple linear regression was performed to explore which variables predicted a significant variation in serum clozapine levels. The predictor variables were selected based on the previous literature and the results of our analysis and included the following clinical variables: daily clozapine dose, sex, smoking status, BMI, and concomitant use of fluvoxamine and/or lamotrigine. The model employed a log transformation of serum clozapine level values as the dependent variable to handle skewed data, similarly to how serum level values were handled in previous predictive models of serum clozapine levels [27,28]. Sex as a predictor was removed, as female sex was collinear with increased BMI (*p* = 0.004) and with being a non-smoker (*p* < 0.001). In addition, the hierarchical procedure demonstrated that the addition of sex as a predictor did not contribute significantly to log-transformed serum clozapine levels (*p* = 0.87). The resultant model explained a significant amount of variance (43.6%) in serum clozapine levels (Table 4). The use of fluvoxamine was the most important predictor of increased serum clozapine levels and predicted an increase of approximately 455.9%. The second most important predictor was daily dose, as each 1 mg increase in dose predicted a 0.23% increase in serum clozapine levels. In addition, being a non-smoker predicted a 51.4% increase in serum clozapine levels, whereas the use of lamotrigine predicted a significant increase in serum clozapine levels of approximately 112.8%. BMI was the last predictor in the model, with each single unit increment in BMI predicting a 1.86% increase in serum clozapine levels. The model equation for predicting serum clozapine levels in our sample of Middle Eastern patients was as follows: log_10_ (clozapine level) = 2.135 + (0.745 × fluvoxamine use) + (0.001 × clozapine dose) + (0.180 × being a non-smoker) + (0.328 × lamotrigine use) + (0.008 × BMI), where categorical clinical variables were substituted by 1 if present.

**Table 4.** Multiple linear regression of serum clozapine levels (N = 94)

| Clinical predictor | B | 95% CI B | beta | t | p |
| --- | --- | --- | --- | --- | --- |
| (Constant) | 2.135 | (1.879–2.391) |  | 16.57 | <0.001 |
| Use of fluvoxamine | 0.745 | (0.516, 0.975) | 0.511 | 6.46 | <0.001 |
| Clozapine daily dose | 0.001 | (0.001, 0.001) | 0.323 | 4.08 | <0.001 |
| Non-smoker | 0.180 | (0.066, 0.293) | 0.249 | 3.14 | 0.002 |
| Use of lamotrigine | 0.328 | (0.064, 0.592) | 0.196 | 2.47 | 0.016 |
| Body mass index | 0.008 | (0.001, 0.015) | 0.177 | 2.22 | 0.029 |

Dependent variable: $\log_{10}$ (serum clozapine levels)
Model fit: $F(6, 86) = 15.246$ , $p < 0.001$ , $R^2 = 0.467$ , $R^2_{\text{Adjusted}} = 0.436$
CI: confidence interval

## Discussion

To the best of our knowledge, this is currently the largest study investigating serum clozapine levels in Middle Eastern patients and the first study on this research question in more than two decades [21]. Similar to those of previous studies, our results demonstrated a linear association between the daily dose of clozapine and serum levels [13,20,23]. The study results are also consistent with our clinical observation that the association between clozapine dose and serum levels in patients of Middle Eastern ethnicity resembles the dose-level association found in East Asians as opposed to European Caucasians [13,18,19,22]. Moreover, women had significantly higher serum clozapine levels in our sample than men, as they did in other populations [20,27,28]. However, our correlational analysis suggested that the factor of female sex was confounded by smoking status and BMI. We observed that an increase in BMI predicted increased serum clozapine levels, which is in agreement with a previous study suggesting a positive correlation between weight and serum levels of clozapine [27]. Smoking status was strongly associated with clozapine levels in our sample, which is consistent with numerous reports demonstrating that smoking cigarettes is associated with decreased serum clozapine levels [27,29,30]. However, this association was only observed in our male patients, as there was an inadequate number of female smokers in our sample for statistical analysis. It is well established that the use of fluvoxamine increases serum clozapine levels significantly [31], and in our sample, the use of fluvoxamine was the most significant predictor of higher serum clozapine levels. Furthermore, we observed that the use of lamotrigine also predicted a significant increase in clozapine serum levels. However, this effect has only been documented in one case report [32], and very few patients were using lamotrigine in our sample. Other researchers have concluded that lamotrigine does not significantly affect clozapine metabolism [33]. Using our predictive model detailed in Table 4, we calculated the following as the maximum clozapine dose to avoid exceeding a serum level of 650 ng/mL for our Middle Eastern patients: 300, 250, or 225 mg daily for non-smokers with a BMI of 25, 30, or 35 kg/m^2^, respectively, and 475, 425, or 400 mg daily for smokers with a BMI of 25, 30, or 35 kg/m^2^, respectively. Maximum doses could not be calculated for patients concomitantly receiving fluvoxamine or lamotrigine because the model predicted levels above the optimal range (higher than 650 ng/mL) if the patient was receiving any one of these medications, especially if they were non-smokers. It is important to note that these calculated dose values are based on approximate definitions of the optimal range of clozapine serum levels, and the small sample size does not allow these findings to be a substitute for TDM in clinical practice.

Since the association between serum clozapine levels and clinical response is believed to be curvilinear [34], clinicians treating patients of Middle Eastern descent are encouraged to utilise TDM in clozapine therapy to ensure treatment success. If TDM is unavailable, our predictive model may serve as a preliminary guide to mitigate the reluctance of clinicians to prescribe clozapine to Middle Eastern patients. Our findings highlight both the benefits of studying psychotropic medications in different ethnic groups and the risk of neglecting ethnic differences in biomedical research at large [35].

This study has some limitations, including small sample size, retrospective design, and reliance on clinical notes in electronic medical records. The collected data did not include possible confounding factors such as dietary habits, which may have affected drug metabolism. In determining our sample as Middle Eastern, we relied on nationality and family or tribal affiliations but did not conduct any testing to confirm ethnicity. Moreover, we could not collect data on possible deviation from the standardised time of sampling by the laboratory personnel. Finally, our sample was heterogeneous with patients having different diagnoses and treatment regimens.

## Conclusions

Our results provide a naturalistic examination of clozapine TDM, which reflects daily practice in the Middle eastern country of Saudi Arabia. The findings demonstrate the relationship between clozapine dose and serum levels in Middle Eastern patients, which can be compared with findings of other reports in the literature. In addition, our results include a predictive model to estimate serum clozapine levels with common variables, which can be clinically useful notwithstanding its limitations. These findings can be helpful to clinicians prescribing clozapine to Middle Easterners but should not be a substitute to individualised TDM. Future studies should investigate the association between clozapine dose and serum levels in larger samples of Middle Eastern patients and further explore the association between serum clozapine levels and clinical response in this demographic.

## Data Availability

The datasets analysed in the current study are available from the corresponding author on reasonable request.

## List of Abbreviations

TDM: therapeutic drug monitoring
BMI: body mass index

## Declarations

### Ethics approval and consent to participate

The study was approved by the Institutional Research Board Committee at the College of Medicine at King Saud University (approval reference no. 19/0054/IRB for Research Project No. E-19-4246). The requirement for informed consent was waived by the institutional review board because the study employed a retrospective observational design, and the patients’ data remained confidential.

### Consent for publication

Not applicable.

### Competing interests

The authors declare that they have no competing interests

### Funding

This research did not receive any specific grant from funding agencies in the public, commercial, or not-for-profit sectors.

### Authors’ contributions

A.H. designed the research, supervised data collection and analysis, and reviewed the manuscript; M.A. performed the data collection and analysis, and wrote the manuscript under supervision of the lead author. Both authors have approved of this final manuscript.

## Acknowledgements

We would like to thank Editage (www.editage.com) for English language editing. This study was conducted at King Saud University.

## Authors’ information

1. Ahmed Hassab Errasoul, MBBS, DCP, MMedSc, MRCPsych, CCST Consultant Adult Psychiatrist
2. Mohammed A. Alarabi, MBBS Senior Registrar Psychiatrist Clozapine Clinic, Department of Psychiatry, College of Medicine, King Saud University, Riyadh 11472, Saudi Arabia

## Notes

### Competing Interest Statement

The authors have declared no competing interest.

### Author Declarations

The study was approved by the Institutional Research Board Committee at the College of Medicine at King Saud University (approval reference no. 19/0054/IRB for Research Project No. E-19-4246). The requirement for informed consent was waived by the institutional review board because the study employed a retrospective observational design, and patients data remained confidential. The authors assert that all procedures contributing to this work comply with the ethical standards of the relevant national and institutional committee on human experimentation with the Declaration of Helsinki of 1975, as revised in 2008. The authors also followed the ICMJE guidelines in the composition of this manuscript.

### Summary of Updates

This version is updated to match the latest version of the manuscript submitted for peer-review. The changes include formatting and paraphrasing to fit the guidelines and word limit of the target journal. There are no differences in the study results or interpretation.

## References

1. Leucht S, Cipriani A, Spineli L, Mavridis D, Örey D, Richter F, et al. Comparative efficacy and tolerability of 15 antipsychotic drugs in schizophrenia: A multiple-treatments meta-analysis. Lancet. 2013;382:951–62.

2. Siskind D, McCartney L, Goldschlager R, Kisely S. Clozapine v. first- and second-generation antipsychotics in treatment-refractory schizophrenia: Systematic review and meta-analysis. Br J Psychiatry. 2016;209:385–92.

3. Huhn M, Nikolakopoulou A, Schneider-Thoma J, Krause M, Samara M, Peter N, et al. Comparative efficacy and tolerability of 32 oral antipsychotics for the acute treatment of adults with multi-episode schizophrenia: A systematic review and network meta-analysis. Lancet. 2019 Jul 11 Cited 2019 Aug 29;394:939–51. http://www.ncbi.nlm.nih.gov/pubmed/31303314.

4. Warnez S, Alessi-Severini S. Clozapine: A review of clinical practice guidelines and prescribing trends. BMC Psychiatry. 2014;14:102.

5. Howes OD, Vergunst F, Gee S, McGuire P, Kapur S, Taylor D. Adherence to treatment guidelines in clinical practice: Study of antipsychotic treatment prior to clozapine initiation. Br J Psychiatry. 2012 Dec 2 Cited 2018 Nov 17;201:481–5. http://www.ncbi.nlm.nih.gov/pubmed/22955007.

6. Alosaimi FD, Alhabbad A, Abalhassan MF, Fallata EO, Alzain NM, Alassiry MZ, et al. Patterns of psychotropic medication use in inpatient and outpatient psychiatric settings in Saudi Arabia. Neuropsychiatr Dis Treat. 2016;12:897–907.

7. Krivoy A, Malka L, Fischel T, Weizman A, Valevski A. Predictors of clozapine discontinuation in patients with schizophrenia. Int Clin Psychopharmacol. 2011;26:311–5.

8. Mustafa FA, Burke JG, Abukmeil SS, Scanlon JJ, Cox M. Schizophrenia past clozapine: Reasons for clozapine discontinuation, mortality, and alternative antipsychotic prescribing. Pharmacopsychiatry. 2014 Nov 6 Cited 2018 Nov 17;45:11–4. http://www.ncbi.nlm.nih.gov/pubmed/25376977.

9. Ismail D, Tounsi K, Zolezzi M, Eltorki Y. A qualitative exploration of clozapine prescribing and monitoring practices in the Arabian Gulf countries. Asian J Psychiatr. 2019;39:93–7.

10. Kang JS, Lee MH. Overview of therapeutic drug monitoring. Korean J Intern Med. 2009;24:1–10.

11. Miller DD, Fleming F, Holman TL, Perry PJ. Plasma clozapine concentrations as a predictor of clinical response: A follow-up study. J Clin Psychiatry. 1994 Sep Cited 2018 Nov 17;55;Suppl B:117–21. http://www.ncbi.nlm.nih.gov/pubmed/7961554.

12. Kronig MH, Munne RA, Szymanski S, Safferman AZ, Pollack S, Cooper T, et al. Plasma clozapine levels and clinical response for treatment-refractory schizophrenic patients. Am J Psychiatry. 1995 Feb Cited 2018 Nov 18;152:179–82. http://www.ncbi.nlm.nih.gov/pubmed/7840349.

13. VanderZwaag C, McGee M, McEvoy JP, Freudenreich O, Wilson WH, Cooper TB. Response of patients with treatment-refractory schizophrenia to clozapine within three serum level ranges. Am J Psychiatry. 1996;153:1579–84.

14. Bell R, McLaren A, Galanos J, Copolov D. The clinical use of plasma clozapine levels. Aust N Z J Psychiatry. 1998 Aug 26 Cited 2018 Nov 17;32:567–74. http://journals.sagepub.com/doi/10.3109/00048679809068332.

15. Spina E, Avenoso A, Facciolà G, Scordo MG, Ancione M, Madia AG, et al. Relationship between plasma concentrations of clozapine and norclozapine and therapeutic response in patients with schizophrenia resistant to conventional neuroleptics. Psychopharmacol (Berl). 2000 Jan Cited 2018 Nov 17;148:83–9. http://www.ncbi.nlm.nih.gov/pubmed/10663421.

16. Nielsen J, Damkier P, Lublin H, Taylor D. Optimizing clozapine treatment. Acta Psychiatr Scand. 2011 Jun Cited 2018 Nov 18;123:411–22. http://www.ncbi.nlm.nih.gov/pubmed/21534935.

17. Remington G, Agid O, Foussias G, Ferguson L, McDonald K, Powell V. Clozapine and therapeutic drug monitoring: Is there sufficient evidence for an upper threshold? Psychopharmacol (Berl). 2013 Feb 22 Cited 2018 Nov 18;225:505–18. http://www.ncbi.nlm.nih.gov/pubmed/23179967.

18. Couchman L, Morgan PE, Spencer EP, Flanagan RJ. Plasma clozapine, norclozapine, and the clozapine:norclozapine ratio in relation to prescribed dose and other factors: Data from a therapeutic drug monitoring service, 1993-2007. Ther Drug Monit. 2010;32:438–47.

19. Ng CH, Chong SA, Lambert T, Fan A, Hackett LP, Mahendran R, et al. An inter-ethnic comparison study of clozapine dosage, clinical response and plasma levels. Int Clin Psychopharmacol. 2005 May Cited 2018 Nov 19;20:163–8. http://www.ncbi.nlm.nih.gov/pubmed/15812267.

20. Wohkittel C, Gerlach M, Taurines R, Wewetzer C, Unterecker S, Burger R, et al. Relationship between clozapine dose, serum concentration, and clinical outcome in children and adolescents in clinical practice. J Neural Transm (Vienna). 2016;123:1021–31.

21. Hussein R, Gad A, Raines DA, Chaleby K, Al-Rawithi S, El-Yazigi A. Steady-state pharmacokinetics of clozapine in refractory schizophrenic Saudi Arabian patients. Pharm Pharmacol Commun. 1999;5:473–8.

22. Chang WH, Lin SK, Lane HY, Hu WH, Jann MW, Lin HN. Clozapine dosages and plasma drug concentrations. J Formos Med Assoc. 1997 Aug Cited 2018 Nov 19;96:599–605. http://www.ncbi.nlm.nih.gov/pubmed/9290269.

23. Rajkumar AP, Poonkuzhali B, Kuruvilla A, Jacob M, Jacob KS. Clinical predictors of serum clozapine levels in patients with treatment-resistant schizophrenia. Int Clin Psychopharmacol. 2013;28:50–6.

24. Jakobsen MI, Larsen JR, Svensson CK, Johansen SS, Linnet K, Nielsen J, et al. The significance of sampling time in therapeutic drug monitoring of clozapine. Acta Psychiatr Scand. 2017 Feb Cited 2018 Nov 19;135:159–69. http://www.ncbi.nlm.nih.gov/pubmed/27922183.

25. Hiemke C, Baumann P, Bergemann N, Conca A, Dietmaier O, Egberts K, et al. AGNP consensus guidelines for therapeutic drug monitoring in psychiatry: Update 2011. Pharmacopsychiatry. 2011;44:195–235.

26. IBM SPSS Inc. SPSS Statistics for Windows. IBM Corp Released 2012; 2012;version 20. p. 1–8.

27. Rostami-Hodjegan A, Amin AM, Spencer EP, Lennard MS, Tucker GT, Flanagan RJ. Influence of dose, cigarette smoking, age, sex, and metabolic activity on plasma clozapine concentrations: A predictive model and nomograms to aid clozapine dose adjustment and to assess compliance in individual patients. J Clin Psychopharmacol. 2004 Feb Cited 2018 Nov 17;24:70–8. http://www.ncbi.nlm.nih.gov/pubmed/14709950.

28. Lane HY, Chang YC, Chang WH, Lin SK, Tseng YT Te, Jann MW. Effects of gender and age on plasma levels of clozapine and its metabolites: Analyzed by critical statistics. J Clin Psychiatry. 1999;60:36–40.

29. Meyer JM. Individual changes in clozapine levels after smoking cessation: Results and a predictive model. J Clin Psychopharmacol. 2001;21:569–74.

30. Bersani FS, Capra E, Minichino A, Pannese R, Girardi N, Marini I, et al. Factors affecting interindividual differences in clozapine response: A review and case report. Hum Psychopharmacol. 2011 Cited 2018 Nov 19;26:177–87. https://pdfs.semanticscholar.org/2cff/4b1c47fa0d9d950ae54cbf36d30bd1a71784.pdf.

31. Koponen HJ, Leinonen E, Lepola U. Fluvoxamine increases the clozapine serum levels significantly. Eur Neuropsychopharmacol. 1996 Mar 1;6:69–71.

32. Kossen M, Selten JP, Kahn RS. Elevated clozapine plasma level with lamotrigine (3). Am J Psychiatry. 2001;158:1930.

33. Spina E, D’Arrigo C, Migliardi G, Santoro V, Muscatello MR, Micò U, et al. Effect of adjunctive lamotrigine treatment on the plasma concentrations of clozapine, risperidone and olanzapine in patients with schizophrenia or bipolar disorder. Ther Drug Monit. 2006 Oct;28:599–602.

34. Liu HC, Chang WH, Wei FC, Lin SK, Lin SK, Jann MW. Monitoring of plasma clozapine levels and its metabolites in refractory schizophrenic patients. Ther Drug Monit. 1996;18:200–7.

35. Risch N, Burchard E, Ziv E, Tang H. Categorization of humans in biomedical research: Genes, race and disease. Genome Biol. 2002;3:comment2007.

